# Prevalence and determinants of rheumatic heart disease among school-going children in Dhanusha district, southern Nepal: a cross-sectional echocardiographic screening study

**DOI:** 10.64898/2026.05.15.26353362

**Authors:** Prakash Raj Regmi, Urmila Shakya, Satya Narayan Suwal, Ram Kishore Shah, Rajesh Shah, Prem Raj Baidhya, Ashish Tamang, Shyam Thapa

## Abstract

Rheumatic heart disease (RHD) is a leading preventable cause of cardiac death in children in low and middle-income countries. Nepal’s epidemiological data come mainly from auscultation surveys that miss subclinical disease, and no echocardiographic screening study had been conducted in Dhanusha district, a densely populated, low-income region in southern Nepal. We aimed to determine the prevalence of borderline and definite RHD among school children (6–16 years) in Dhanusha using the 2012 World Heart Federation (WHF) echocardiographic criteria, identify independent predictors, and quantify school-level clustering via the intraclass correlation coefficient (ICC). In a cross-sectional study (January 2023–December 2024), we screened 4,536 children from 8 public schools selected by four-stage cluster sampling. RHD was classified by WHF 2012 criteria; predictors were identified using random-effects logistic regression with school as random intercept. Ethical approval was from the Nepal Health Research Council (Protocol No. 155/2023). Overall prevalence of borderline or definite RHD was 18.7 per 1,000 (95% CI 15.1–23.0); definite RHD was 6.8 per 1,000 (95% CI 4.7–9.7) and borderline RHD 11.9 per 1,000 (95% CI 9.0–15.5). Prevalence was higher in girls (23.3 per 1,000) than boys (13.6 per 1,000; P=0.02), with the peak in girls aged 10–14 years (26.0 per 1,000). Subclinical disease accounted for 64.7% of cases; auscultation sensitivity was 35.3%. Mitral valve involvement predominated. Female sex was the sole independent predictor (OR 1.60, 95% CI 1.02–2.53; P=0.043). The school-level ICC was 0.19 (95% CI 0.07–0.44; P<0.001), giving a design effect of ≈109. The echocardiographic RHD burden in Dhanusha (18.7 per 1,000) is the highest documented in Nepal. Two-thirds of cases are subclinical. Female sex and school attended explain a similar amount of variance in RHD risk, supporting school-targeted screening and informing sample size planning for future cluster-based surveillance.

## Introduction

Rheumatic heart disease (RHD) is the most common acquired cardiac condition among children and young adults across low and middle-income countries. It arises as an autoimmune sequel to group A *Streptococcus* (GAS) pharyngitis that triggers acute rheumatic fever (ARF) and, through recurrent episodes, causes progressive valvular damage, predominantly affecting the mitral valve. An estimated 40.5 million people live with RHD globally and it accounts for more than 300,000 deaths each year, with the overwhelming majority of that burden concentrated in South and Southeast Asia, sub-Saharan Africa and Oceania [1–3]. Although Nepal has acknowledged RHD as a public health problem for decades, reliable echocardiographic prevalence data are still lacking [4].

Auscultation-based surveillance, the dominant approach in most Nepalese studies, systematically misses subclinical disease. Echocardiographic screening consistently reveals that subclinical RHD is five to eight times more prevalent than clinically manifest disease [5,6], and the sensitivity of cardiac auscultation for RHD detection rarely exceeds 20–30% even in experienced hands [7]. The 2012 World Heart Federation (WHF) echocardiographic criteria provide the current gold standard for distinguishing borderline from definite disease [8], enabling early identification before irreversible valve damage occurs and secondary antibiotic prophylaxis can still prevent progression [5].

The only large echocardiographic screening study in Nepal, conducted in Sunsari district, eastern Nepal, documented a prevalence of 10.2 per 1,000 (95% CI 7.5–13.0) using WHF 2012 criteria [9]. Earlier auscultation-based surveys from the Kathmandu Valley and hill districts reported 0.9–7.2 per 1,000 [10–12]. Dhanusha district in the southern Terai had never been the subject of an echocardiographic RHD survey, despite characteristics that suggest potentially high burden: elevated population density, a predominantly agricultural and remittance-dependent economy, limited access to specialist healthcare and a diverse Madhesi population in which conditions favouring GAS transmission are widely prevalent. Beyond prevalence estimation, no prior Nepalese study had quantified the ICC for school-level RHD clustering, a parameter essential both for understanding whether structural school-level factors contribute independently to disease risk and for powering future cluster-based surveillance studies correctly.

The objectives of this study were: (1) to determine the prevalence of borderline and definite RHD among school-going children aged 6–16 years in Dhanusha district using WHF 2012 echocardiographic criteria; (2) to identify independent individual-level predictors of RHD; and (3) to estimate the ICC for school-level clustering and its implications for future study design.

## Methods and Materials

We conducted a cross-sectional, school-based echocardiographic screening study in Dhanusha district, Madhesh Province, Nepal, between January 2023 and December 2024. The study adhered to the Strengthening the Reporting of Observational Studies in Epidemiology (STROBE) guidelines [13]. A completed STROBE checklist is provided as a supplementary file (S1 Checklist). The study is part of a Nepal Heart Foundation programme that has conducted similar echocardiographic surveys across more than 25 districts in Nepal.

Ethical approval was obtained from the Nepal Health Research Council (NHRC Protocol No. 155/2023; approved 13 April 2023). Written informed consent was obtained from parents or guardians, and assent was obtained from children (verbal for ages 7–11 years; written for ages 12–16 years). For illiterate caregivers, thumbprint consent witnessed by a third party was accepted.

Parents and guardians were informed about the screening through a written notice sent home with each child one week prior to the scheduled screening date, followed by an in-person explanation by study staff on the day of screening before consent was obtained.

Dhanusha district comprises one sub-metropolitan city (Janakpurdham), 11 urban municipalities and 6 rural municipalities across 169 wards, with a total population of approximately 873,274 (2021 census). The economy is dominated by smallholder agriculture and labour migration remittances. The population is ethnically diverse, primarily Madhesi communities (Dalit, Yadav, Muslim, Teli, Dhanuk and Janajati) with historically limited access to specialist healthcare. The study was conducted in partnership with the Shahid Gangalal National Heart Center (SGNHC) branch hospital in Janakpur.

The study population comprised children aged 6–16 years enrolled in public school grades pre-primary through grade 10 in Dhanusha district. Private schools, which serve approximately 15% of total enrolment from higher-income households, were excluded so that the study would characterise burden in the most socioeconomically vulnerable group. One consequence of this exclusion is that, to the extent that higher income is associated with lower RHD risk, our prevalence estimates may be modestly upward-biased relative to the full district school-going population.

A four-stage cluster sampling strategy was applied. Stage I: Municipalities were selected from the 18 administrative units of Dhanusha, restricted to those with at least two schools spanning grades 1–5, 6–8 and 9–10 (11 of 18 qualified). Using a stratified purposive sampling approach to ensure representation across geographic and socioeconomic strata, eight municipalities were selected. Consequently, the findings may not be fully generalisable to the entire district. Stage II: one ward per municipality was identified based on geographic diversity. Stage III: one large public school was designated as the screening venue per ward. Stage IV: all eligible children present on the scheduled screening day were invited to participate. Each school was visited at least twice to minimise absentee exclusions. The eight participating schools (labelled School A through H) had enrolment numbers ranging from 119 to 1,560 children (median 433).

The minimum required sample size of 4,105 was calculated using the cluster sampling formula n = [DEFF × N × p(1−p)] / [(d²/Z²(N−1)) + p(1−p)], with expected prevalence p = 25 per 1,000, precision d = 2.5 per 1,000, DEFF = 1.5, type I error α = 0.05 and power 80%. Adding 10% for anticipated non-attendance gave a target of 4,515. The enrolled sample of 4,536 exceeded this target.

Inclusion criteria were: all children aged 6–16 years enrolled in grades pre-primary to 10, present at school on the screening day and with written parental consent.

Structured questionnaires administered by trained interviewers in Nepali and Maithili collected: age, sex, school grade, ethnicity, residence, travel time to school, household size, family livelihood, and family economic status (adapted Kuppuswamy scale). The questionnaire was administered to parents or guardians in person at the time of screening. Children were not asked about household economic status. Symptom history in the preceding 12 months was recorded for three items: throat pain or tonsillitis with fever; fever with joint pain in the hands, shoulders, hips or legs; and involuntary jerky movements of the body (chorea).

Cardiac auscultation was performed by a cardiologist in a quiet room with adequate lighting, using a stethoscope. All children subsequently underwent comprehensive transthoracic echocardiography performed by an experienced cardiologist using a portable ultrasound machine (Sonosite M-Turbo with cardiac transducer). Standard imaging views were obtained: parasternal long axis (PLAX), parasternal short axis (PSAX), apical four-chamber (AP4Ch) and apical five-chamber (AP5Ch), with 2D and colour Doppler assessment of the mitral and aortic valves. All children with findings suspicious for RHD on initial echocardiography were referred for confirmatory review by a second senior cardiologist; final diagnosis was consensus-based. All confirmed RHD cases were referred to the Shahid Gangalal National Heart Centre (SGNHC) branch hospital in Janakpur for free counselling, secondary prophylaxis and ongoing follow-up care.

RHD was classified using WHF 2012 echocardiographic criteria [8]. Definite RHD required at least two morphological valve criteria combined with pathological mitral or aortic regurgitation or mitral stenosis, or borderline disease of both the mitral and aortic valves. Borderline RHD required two or more morphological mitral or aortic valve criteria without pathological regurgitation, or pathological regurgitation meeting jet criteria without associated morphological changes. Pathological mitral regurgitation required: maximum jet length ≥2 cm in at least one view; peak velocity ≥3 m/s for one complete envelope; pansystolic jet in at least one view. Pathological aortic regurgitation required: jet length ≥1 cm in at least one view with a pandiastolic jet. Subclinical RHD was defined as echocardiographic RHD without an audible cardiac murmur. Clinically manifest RHD was defined as echocardiographic RHD with an audible murmur.

Baseline characteristics are presented as frequencies and percentages for categorical variables and medians with interquartile ranges for continuous variables. Differences between children with and without RHD were assessed using chi-squared or Fisher’s exact test for categorical variables and the Wilcoxon rank-sum test for age. Prevalence per 1,000 was calculated with exact binomial 95% confidence intervals. Among the 85 RHD-positive cases, echocardiographic and clinical findings were compared by sex using Fisher’s exact test.

To identify independent predictors of RHD, we fitted a random-effects logistic regression model with school as the random intercept. Female sex and age (modelled as a continuous linear variable) were specified a priori as the primary predictors. Results are expressed as odds ratios (ORs) with 95% confidence intervals. The ICC (rho) was estimated; significant school-level clustering was confirmed by a likelihood ratio test comparing the random-effects model against a standard logistic regression. The design effect was computed as DEFF = 1 + (average cluster size − 1) × ICC = 1 + (567 − 1) × 0.19 ≈ 109.

Three pre-specified sensitivity analyses were conducted: (A) addition of a quadratic age term to test the linearity assumption; (B) substitution of categorical age groups (6–9, 10–14 and 15–16 years) for continuous age; and (C) restriction of the outcome to definite RHD only (n=31). All analyses used Stata version 18.0 (StataCorp, College Station, TX, USA). The significance threshold was two-sided α = 0.05.

## Results

### Sample and baseline characteristics

A total of 4,536 children aged 6–16 years from eight public schools across 8 municipalities of Dhanusha district were enrolled and completed echocardiographic screening (Fig 1). Of 4,970 eligible children identified across the 8 schools, 434 were excluded because they fell outside the 6–16 year age range or grade inclusion criteria. The median age of enrolled children was 13 years (IQR 10–15) and 2,402 (53.0%) were female. The sample was predominantly rural (85.2%). The leading ethnic groups were Dalit (15.2%), Muslim (11.8%), Yadav (11.0%) and Janajati (10.7%).

**Figure 1:**
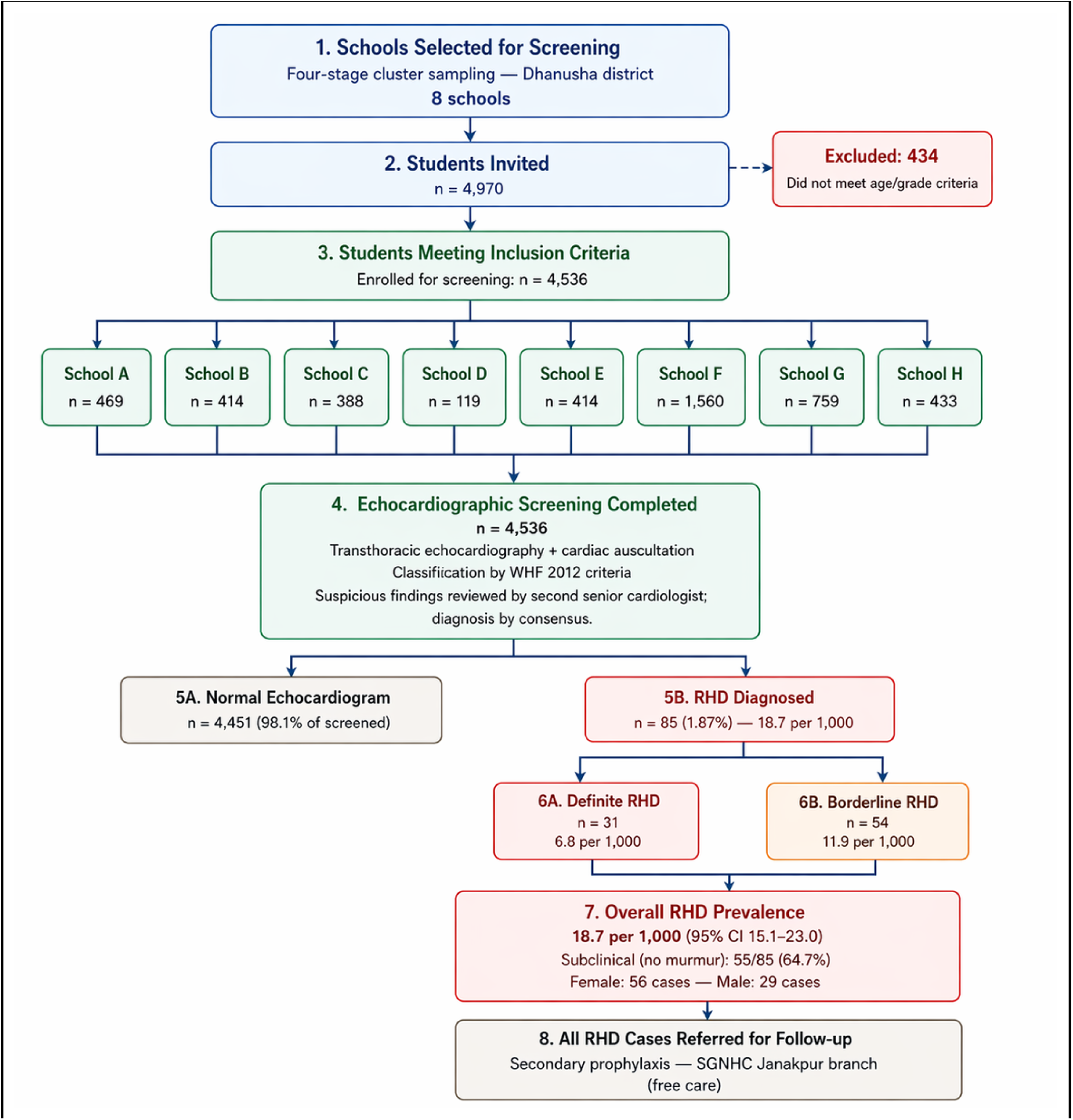
Flow of participant screening and detection of rheumatic heart disease (RHD) among school children in Dhanusha district. *School names have been anonymized with letters A-H

Farming or daily wages was the primary family livelihood for 43.5% of households and employment abroad for 23.8%. Compared with children without RHD, those with RHD were more frequently female (65.9% vs. 52.7%), had a similar median age (13 years in both groups), and were more often from households with low economic status (50.6% vs. 47.8%). No significant differences were observed by ethnicity, residence, or family livelihood. Full baseline characteristics stratified by RHD status are presented in Table 1.

**Table 1.**
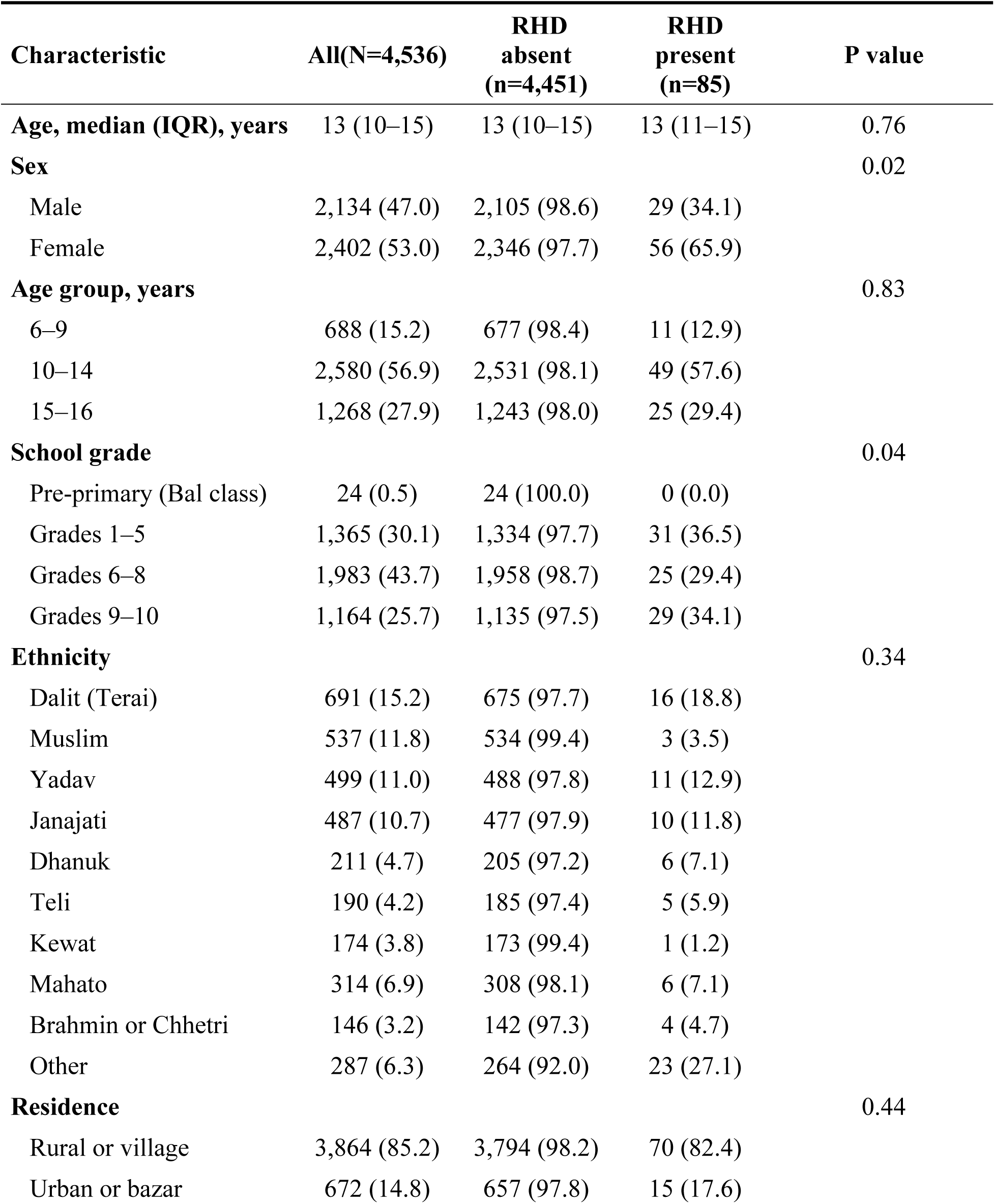
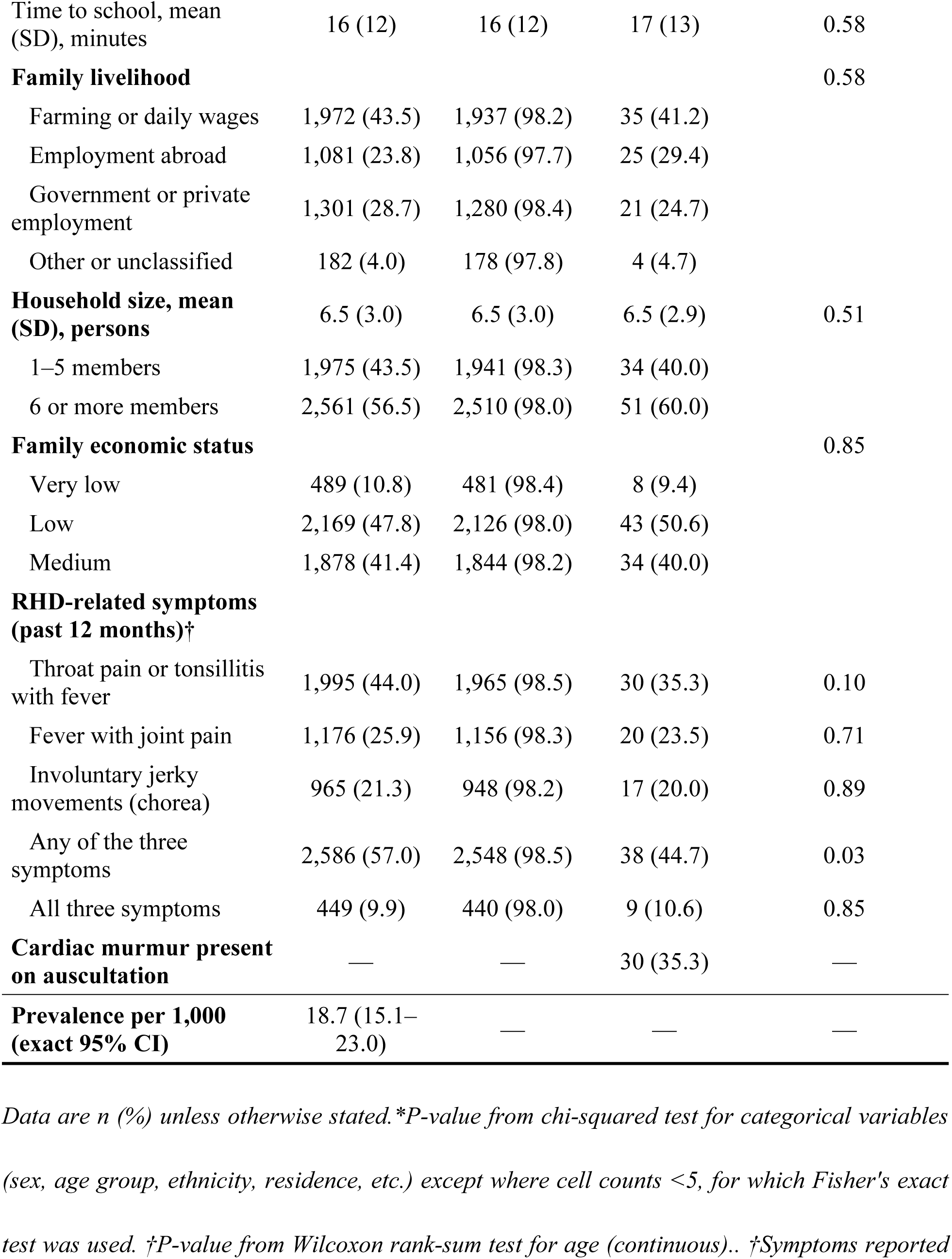
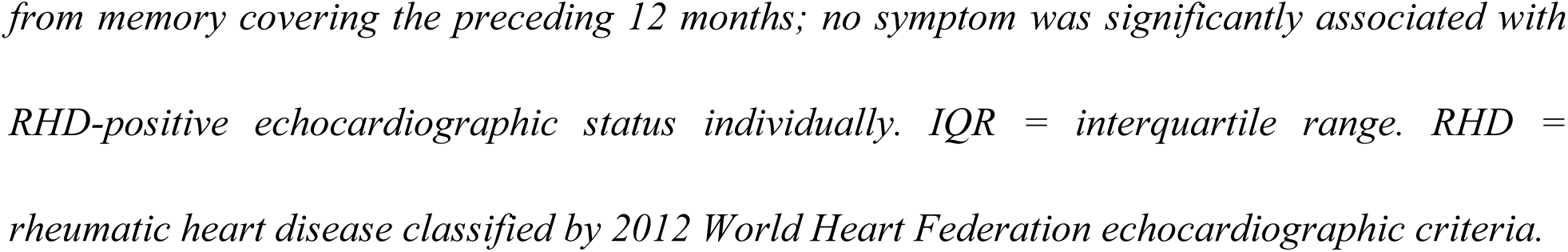
Baseline and sociodemographic characteristics of participants by rheumatic heart disease status (n=4,536)

Eighty-five children (1.87%) met WHF 2012 criteria for borderline or definite RHD: 54 borderline (11.9 per 1,000; 95% CI 9.0–15.5) and 31 definite (6.8 per 1,000; 95% CI 4.7–9.7); overall prevalence 18.7 per 1,000 (95% CI 15.1–23.0). Full prevalence data by type, sex and age group are presented in Table 2.

**Table 2.**
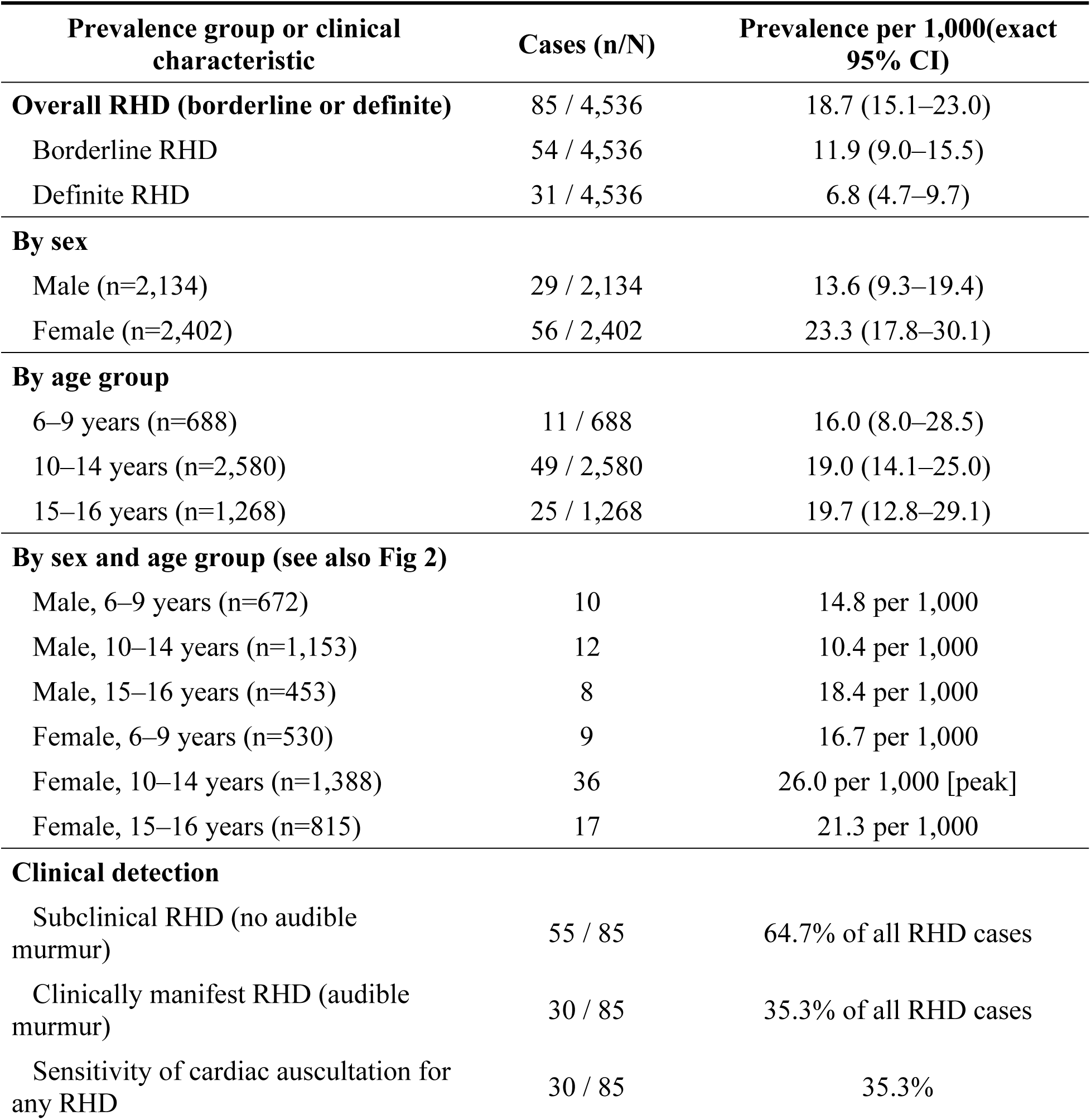
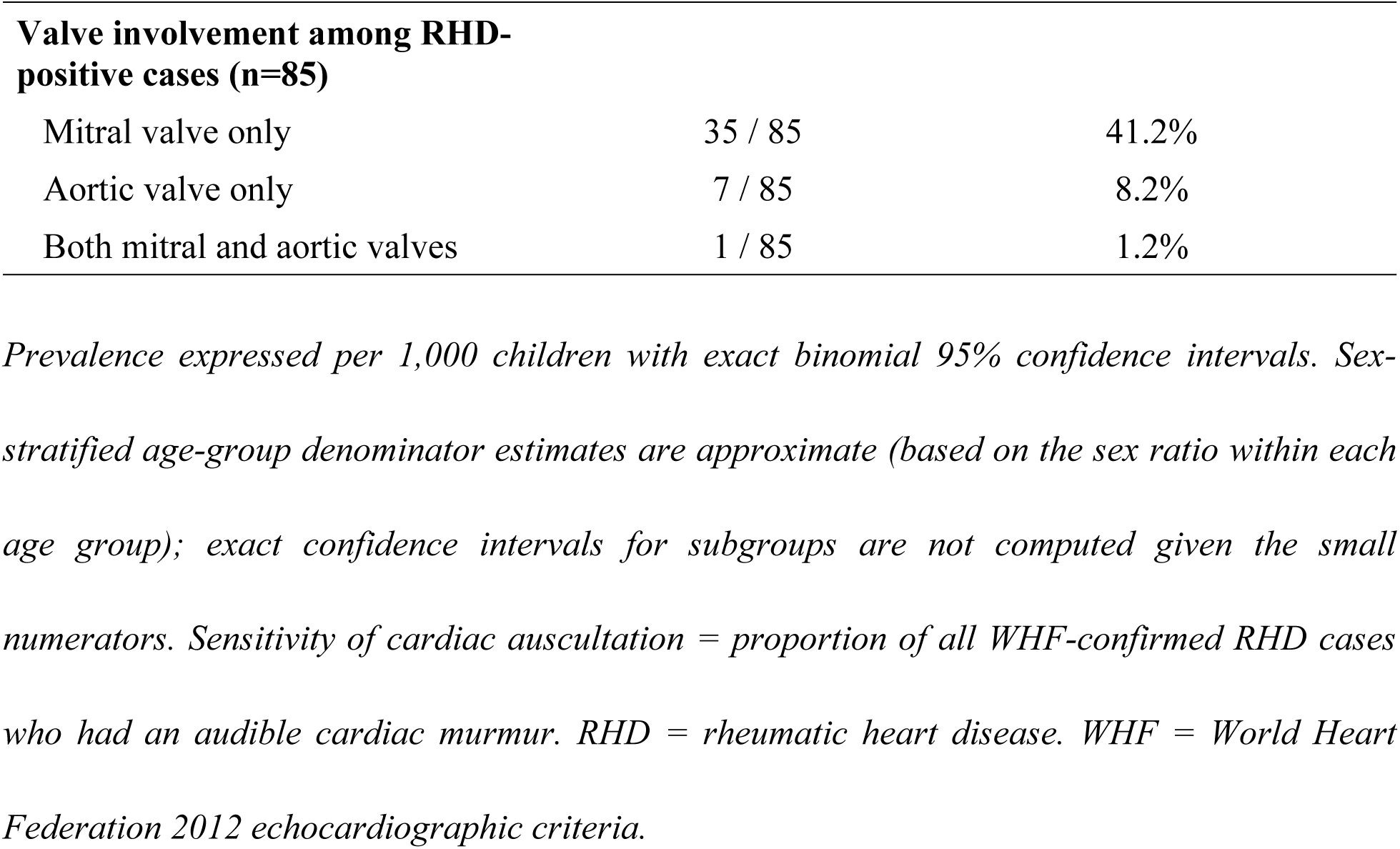
Prevalence of rheumatic heart disease by type, sex and age group; and diagnostic performance of cardiac auscultation (n=4,536)

| **Prevalence group or clinical characteristic** | **Cases (n/N)** | **Prevalence per 1,000(exact 95% CI)** |
| --- | --- | --- |
| **Overall RHD (borderline or definite)** | 85 / 4,536 | 18.7 (15.1–23.0) |
| Borderline RHD | 54 / 4,536 | 11.9 (9.0–15.5) |
| Definite RHD | 31 / 4,536 | 6.8 (4.7–9.7) |
| **By sex** |  |  |
| Male (n=2,134) | 29 / 2,134 | 13.6 (9.3–19.4) |
| Female (n=2,402) | 56 / 2,402 | 23.3 (17.8–30.1) |
| **By age group** |  |  |
| 6–9 years (n=688) | 11 / 688 | 16.0 (8.0–28.5) |
| 10–14 years (n=2,580) | 49 / 2,580 | 19.0 (14.1–25.0) |
| 15–16 years (n=1,268) | 25 / 1,268 | 19.7 (12.8–29.1) |
| **By sex and age group (see also Fig 2)** |  |  |
| Male, 6–9 years (n=672) | 10 | 14.8 per 1,000 |
| Male, 10–14 years (n=1,153) | 12 | 10.4 per 1,000 |
| Male, 15–16 years (n=453) | 8 | 18.4 per 1,000 |
| Female, 6–9 years (n=530) | 9 | 16.7 per 1,000 |
| Female, 10–14 years (n=1,388) | 36 | 26.0 per 1,000 [peak] |
| Female, 15–16 years (n=815) | 17 | 21.3 per 1,000 |
| **Clinical detection** |  |  |
| Subclinical RHD (no audible murmur) | 55 / 85 | 64.7% of all RHD cases |
| Clinically manifest RHD (audible murmur) | 30 / 85 | 35.3% of all RHD cases |
| Sensitivity of cardiac auscultation for any RHD | 30 / 85 | 35.3% |
| **Valve involvement among RHD-positive cases (n=85)** |  |  |
| Mitral valve only | 35 / 85 | 41.2% |
| Aortic valve only | 7 / 85 | 8.2% |
| Both mitral and aortic valves | 1 / 85 | 1.2% |

*Prevalence expressed per 1,000 children with exact binomial 95% confidence intervals. Sex-stratified age-group denominator estimates are approximate (based on the sex ratio within each age group); exact confidence intervals for subgroups are not computed given the small numerators. Sensitivity of cardiac auscultation = proportion of all WHF-confirmed RHD cases who had an audible cardiac murmur. RHD = rheumatic heart disease. WHF = World Heart Federation 2012 echocardiographic criteria.*

Prevalence was substantially higher among girls (23.3 per 1,000; 95% CI 17.8–30.1) than boys (13.6 per 1,000; 95% CI 9.3–19.4; P=0.02). As shown in Fig 2, the sex-stratified age-prevalence patterns differed between males and females. Among boys, prevalence was highest in the 6–9 year group (14.8 per 1,000), fell to 10.4 per 1,000 in the 10–14 year group then rose to 18.4 per 1,000 at 15–16 years. Among girls, prevalence rose from 16.7 per 1,000 at 6–9 years to a peak of 26.0 per 1,000 at 10–14 years, then declined to 21.3 per 1,000 at 15–16 years. The 10–14 year age group in girls thus carried the highest absolute prevalence observed in the entire study.

**Fig 2.**
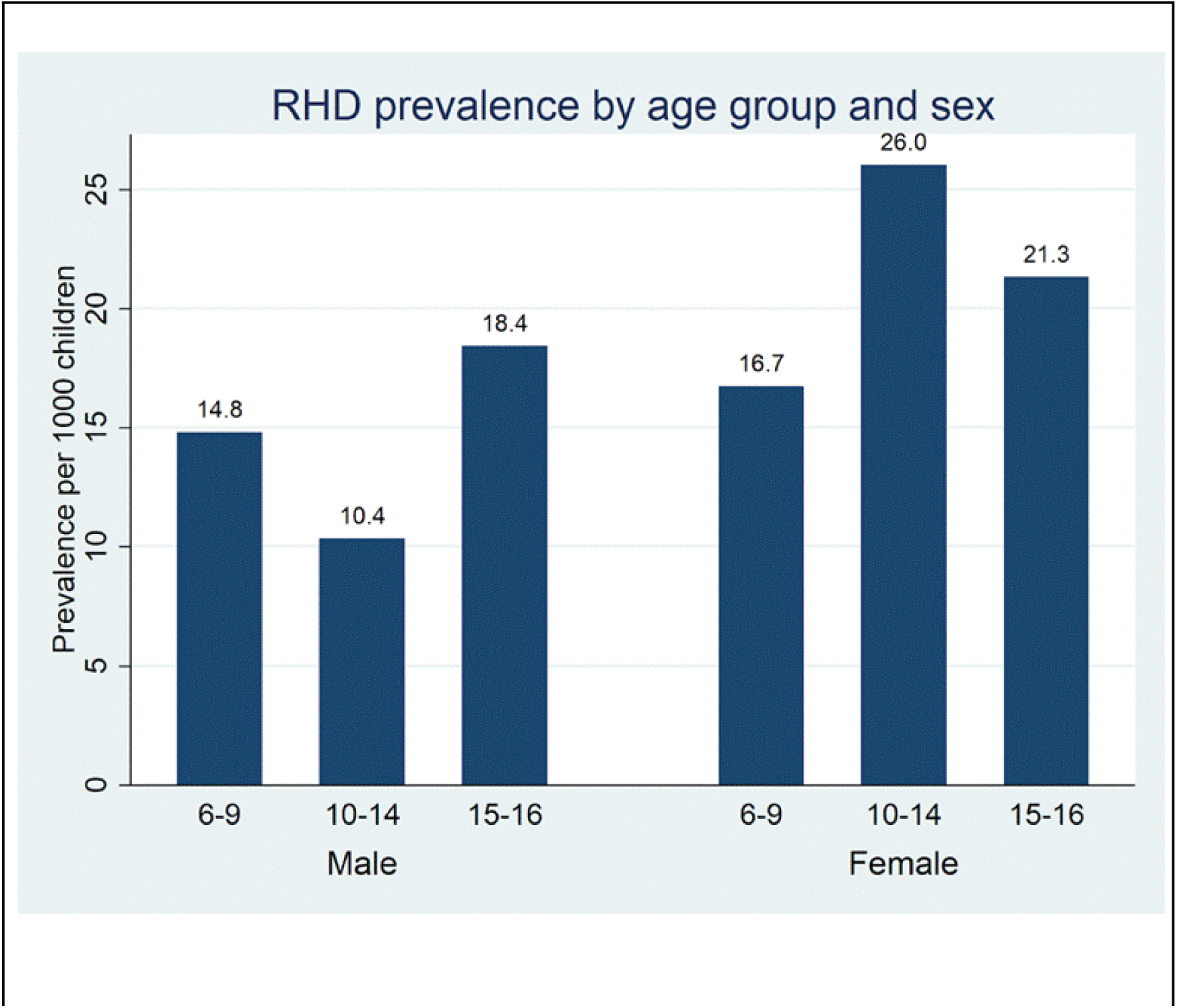
Prevalence of borderline or definite RHD per 1,000 children by age group and sex. Bars show observed prevalence for boys (blue) and girls (coral) across the three age groups (6–9, 10–14 and 15–16 years). Values above bars = prevalence per 1,000 children. Among girls, the 10–14 year group had the highest observed prevalence (26.0 per 1,000); among boys, prevalence dipped at 10–14 years (10.4 per 1,000) before rising at 15–16 (18.4 per 1,000). See Table 2 for exact 95% confidence intervals. RHD = rheumatic heart disease.

No individual RHD-related symptom was significantly associated with RHD-positive status on echocardiography (S2 Table). Throat pain or tonsillitis with fever was the most commonly reported symptom (44.0% of participants), yet children with this history had a lower RHD prevalence (1.5%) than those without it (2.2%; P=0.10). Neither fever with joint pain (P=0.71) nor involuntary jerky movements (P=0.89) was individually associated with RHD. Notably, children who had never experienced any of the three RHD-related symptoms had a higher RHD prevalence (2.4%) than those who reported at least one symptom (1.5%; P=0.03). This finding highlights the predominantly subclinical nature of echocardiographically detected RHD: most affected children had no symptomatic history

Echocardiographic and clinical findings among RHD-positive cases Among the 85 RHD-positive children, mitral valve involvement predominated (Fig 4D). Pathological mitral regurgitation (MR) was present in 76/85 (89.4%) of cases; of those, MR was seen in two or more views in 75/76 (98.7%), peak velocity ≥3 m/s in 72/76 (94.7%), and a pansystolic jet in 72/76 (94.7%). Anterior mitral leaflet thickening ≥3 mm was present in 36/85 (42.4%). Aortic regurgitation was present in 9/85 (10.6%). Detailed echocardiographic findings stratified by sex are presented in S1 Table. No echocardiographic variable differed significantly between males and females among RHD-positive children (all P>0.05). Subclinical disease was the dominant presentation in both sexes: 16/29 (55.2%) of male cases and 39/56 (69.6%) of female cases.

### Multivariable analysis and school-level clustering

In the random-effects logistic regression model with school as random intercept (Table 3 and Fig 3), female sex was independently associated with a 60% higher odds of RHD (OR 1.60; 95% CI 1.02–2.53; P=0.043). The female sex OR was consistent across all three sensitivity analyses: OR 1.61 (P=0.042) with age-squared term; OR 1.61 (P=0.041) with categorical age groups; and OR 1.18 (P=0.651) for definite RHD only (the latter underpowered).

**Fig 3.**
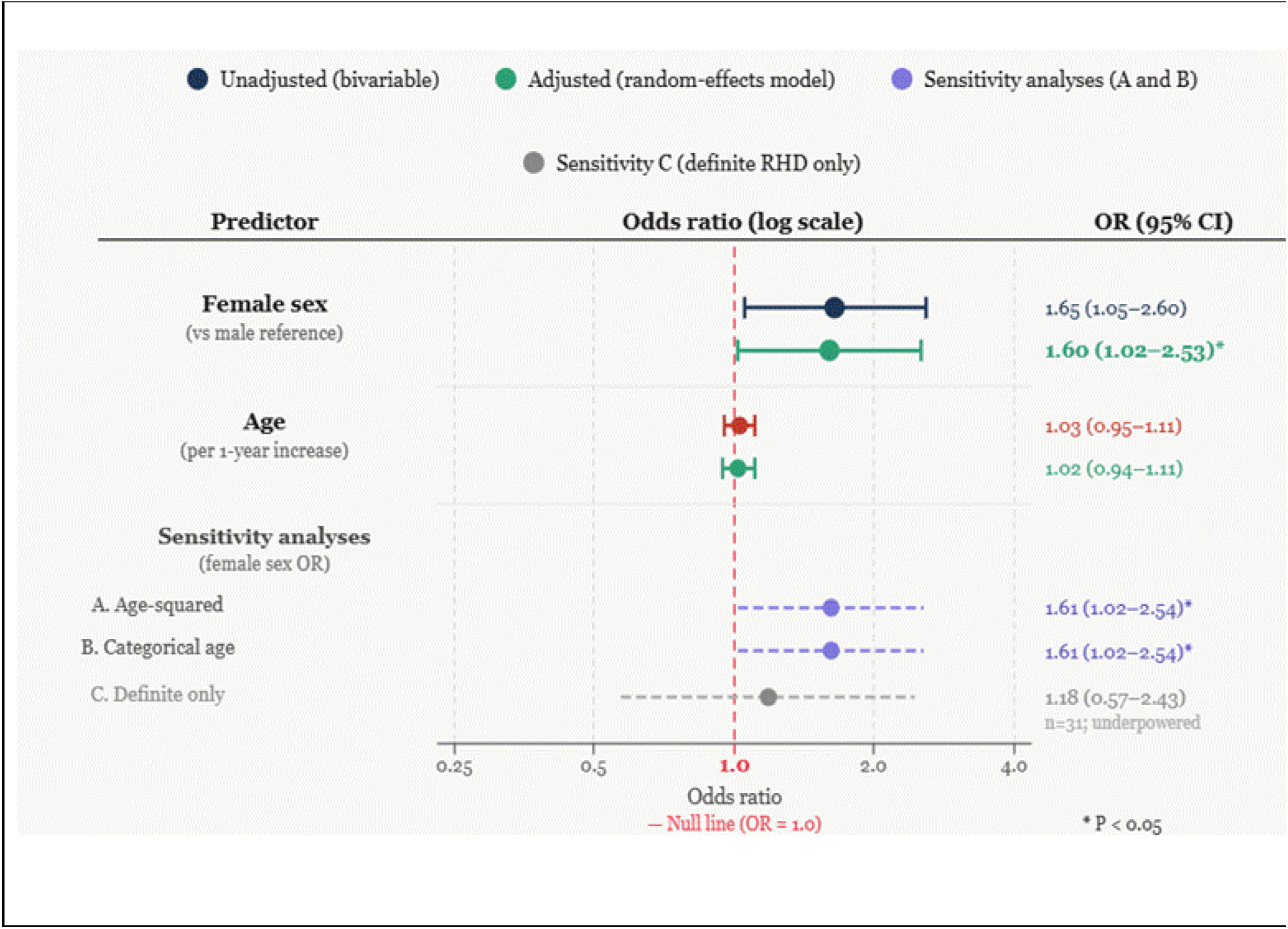
Forest plot of unadjusted (bivariable) and adjusted (multivariable) odds ratios for the two a priori predictors of rheumatic heart disease, Dhanusha district, Nepal, 2023–2024. Adjusted estimates are from the random-effects logistic regression model with school as the random intercept (n=4,536; 85 RHD cases; ICC 0.19; design effect approximately 109; Table 4). Navy circles and lines = unadjusted (bivariable) estimates. Green circles and lines = primary adjusted estimates. Purple dashed lines = sensitivity analyses A (age-squared term added) and B (categorical age groups). Grey dashed line = sensitivity C (definite RHD only; n=31; underpowered). Squares/circles = point estimates. Horizontal lines = 95% confidence intervals. Red dashed vertical line = null value (OR = 1.0). OR = odds ratio. * P < 0.05.

**Table 3.**
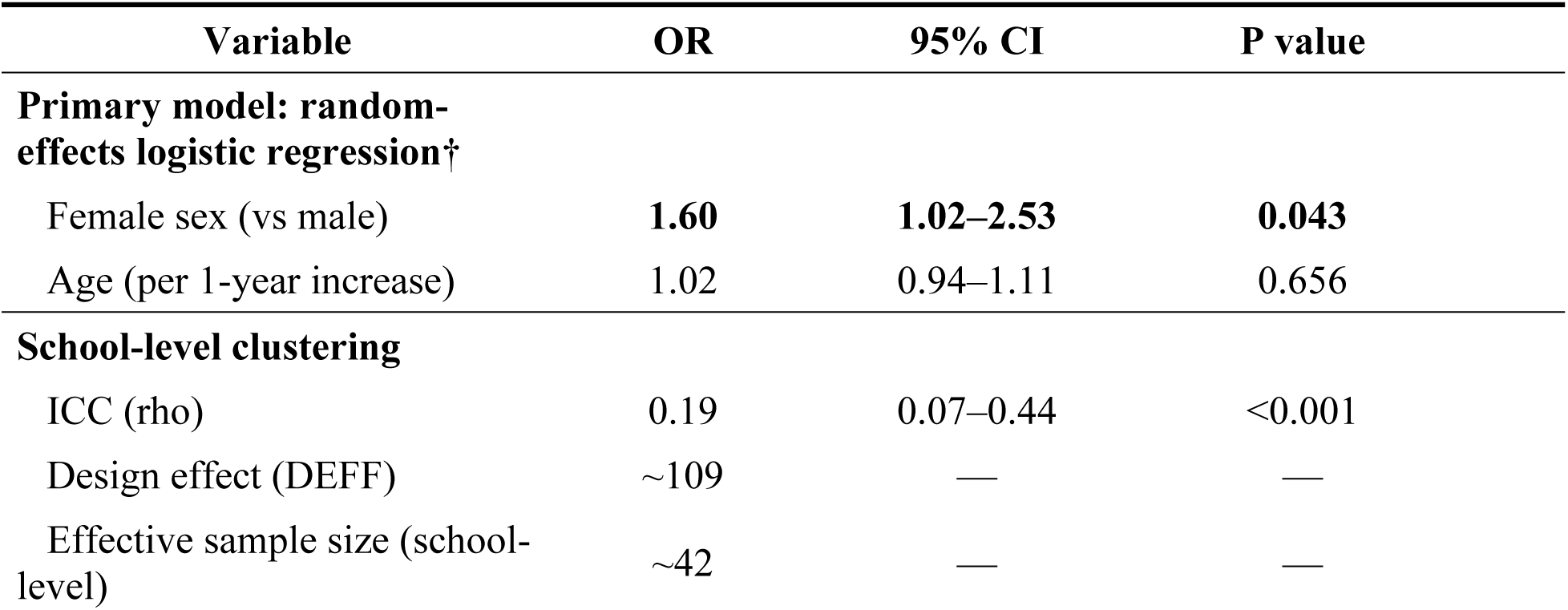
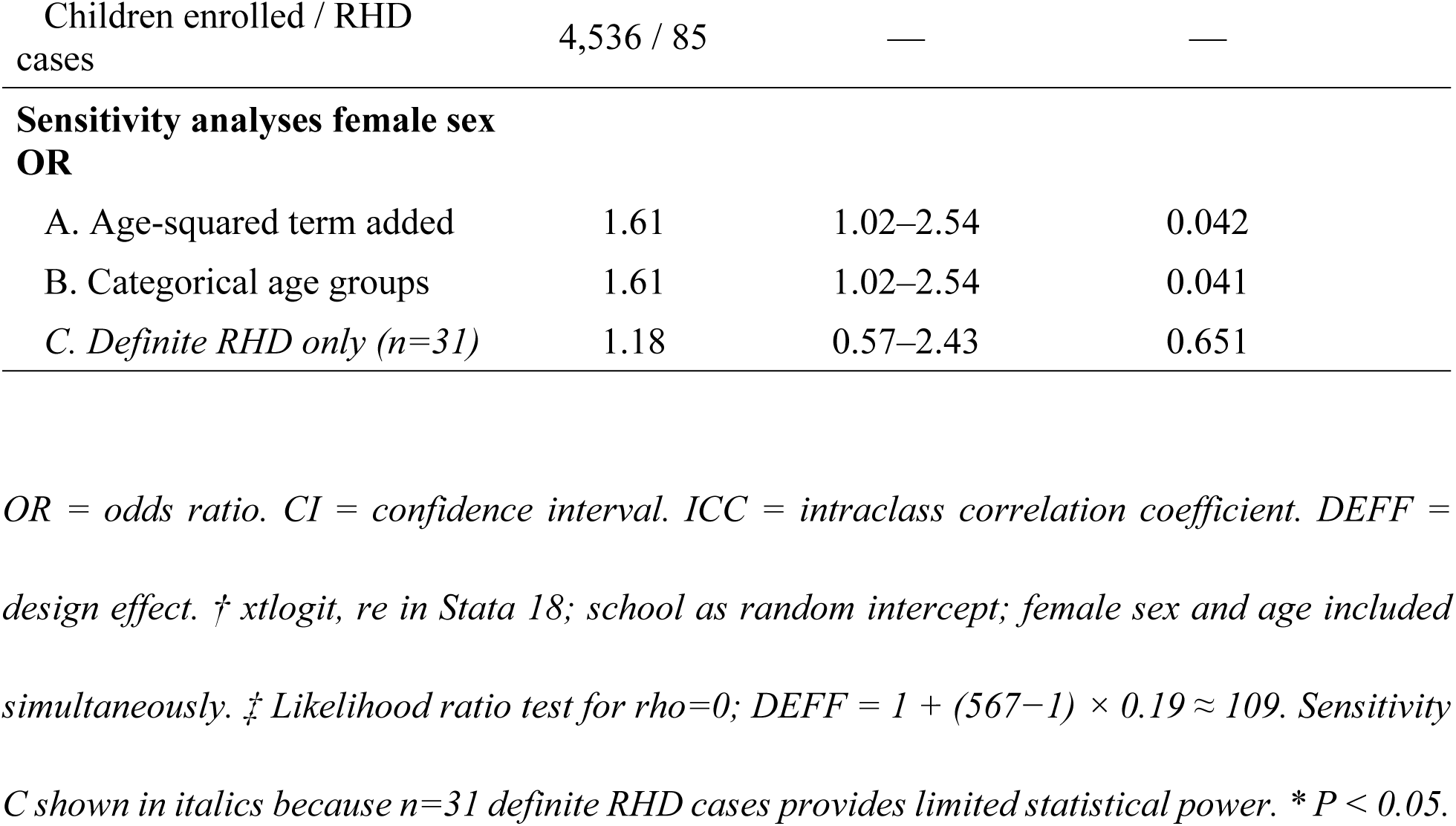
Predictors of rheumatic heart disease: random-effects logistic regression (school as random intercept) and pre-specified sensitivity analyses, (n=4,536)

| Variable | OR | 95% CI | P value |
| --- | --- | --- | --- |
| **Primary model: random-effects logistic regression†** |  |  |  |
| Female sex (vs male) | **1.60** | **1.02–2.53** | **0.043** |
| Age (per 1-year increase) | 1.02 | 0.94–1.11 | 0.656 |
| **School-level clustering** |  |  |  |
| ICC (rho) | 0.19 | 0.07–0.44 | <0.001 |
| Design effect (DEFF) | ~109 | — | — |
| Effective sample size (school-level) | ~42 | — | — |
| Children enrolled / RHD cases | 4,536 / 85 | — | — |
| **Sensitivity analyses female sex OR** |  |  |  |
| A. Age-squared term added | 1.61 | 1.02–2.54 | 0.042 |
| B. Categorical age groups | 1.61 | 1.02–2.54 | 0.041 |
| *C. Definite RHD only (n=31)* | 1.18 | 0.57–2.43 | 0.651 |

*OR = odds ratio. CI = confidence interval. ICC = intraclass correlation coefficient. DEFF = design effect. † xtlogit, re in Stata 18; school as random intercept; female sex and age included simultaneously. ‡ Likelihood ratio test for $\rho=0$ ; $DEFF = 1 + (567-1) \times 0.19 \approx 109$ . Sensitivity C shown in italics because n=31 definite RHD cases provides limited statistical power. \* $P < 0.05$ .*

The ICC was 0.19 (95% CI 0.07–0.44; LR test P<0.001), indicating significant school-level clustering of RHD risk. School attended explained 19% of the total variance in RHD occurrence after accounting for sex and age. The design effect of approximately 109 means that this study of 4,536 children across eight schools provides inferential precision for school-level comparisons equivalent to only about 42 independent observations.

## Discussion

This echocardiographic screening study of 4,536 school-going children aged 6–16 years in Dhanusha district documents an RHD prevalence of 18.7 per 1,000 (borderline + definite, WHF 2012), the highest reported from Nepal to date. This estimate is substantially higher than the 10.2 per 1,000 observed in Sunsari [9] and exceeds the pooled estimate of 10.6 per 1,000 from Nepalese echocardiographic studies [15]. In contrast, studies relying on auscultation followed by confirmatory echocardiography report markedly lower prevalence: 7.32 per 1,000 in rural Jajarkot [16], 2.22 per 1,000 from a large national Nepal Heart Foundation programme [15], and 0.90 per 1,000 in urban Kathmandu schoolchildren [17].

The burden peaks among girls aged 10–14 years at 26.0 per 1,000 (Fig 2) and nearly two-thirds of all RHD cases are subclinical (Fig 4B). Mitral valve involvement predominates and echocardiographic phenotype does not differ significantly by sex among affected children. Female sex was the only independently significant individual-level predictor of RHD (OR 1.60; Fig 3; Table 3), a finding that remained consistent across sensitivity analyses. The observed school-level clustering (ICC 0.19) indicates that a substantial proportion of variation in RHD risk is attributable to the school level, highlighting the potential importance of shared environmental and contextual factors. This has direct implications for the design of future cluster-based surveillance studies and for targeting echocardiographic screening strategies.

**Fig 4.**
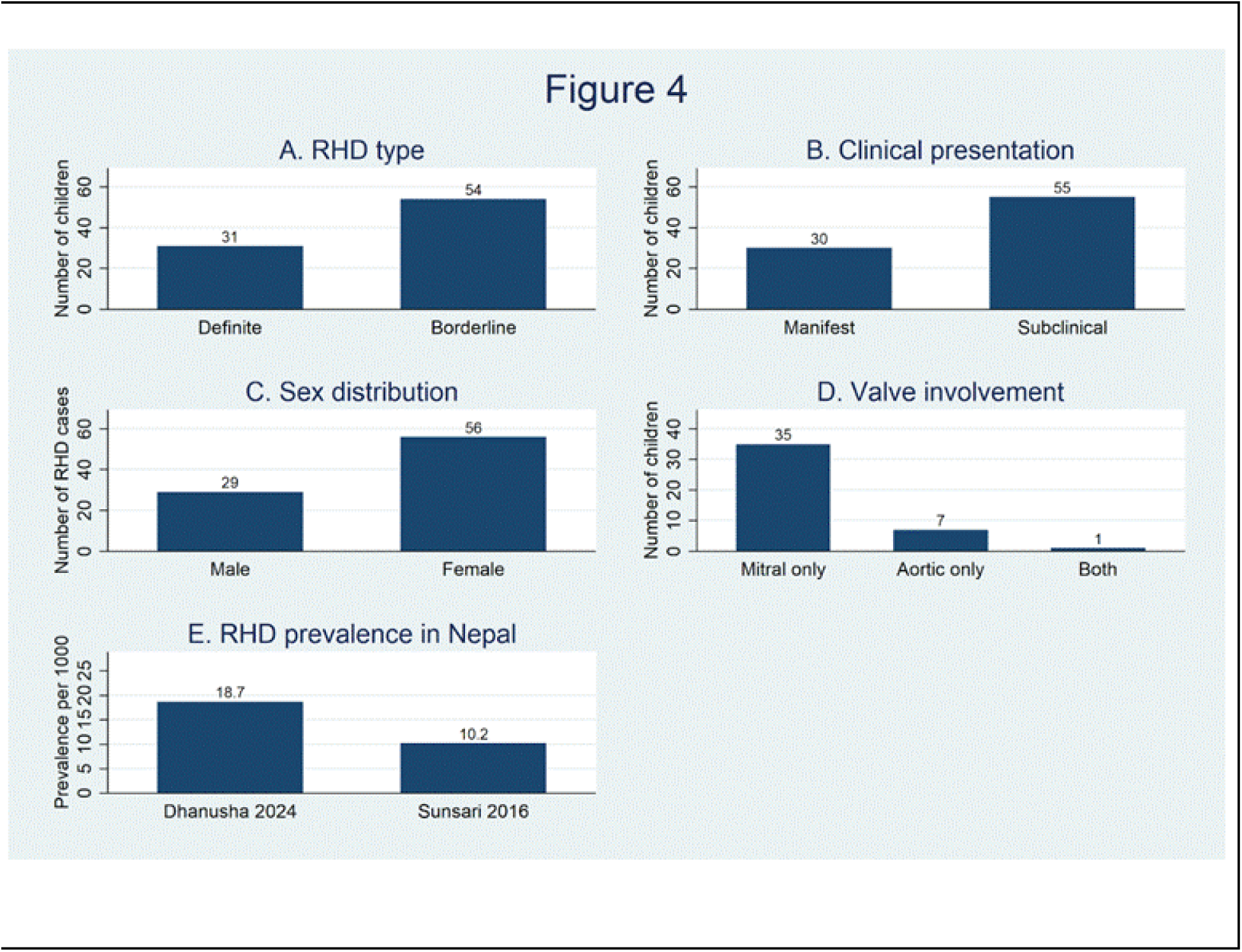
Summary of echocardiographic and clinical findings among the 85 children with rheumatic heart disease, Dhanusha district, Nepal, 2023–2024. Panel A: RHD type (borderline RHD n=54; definite RHD n=31). Panel B: clinical presentation subclinical (no audible murmur; n=55) versus clinically manifest (audible murmur; n=30). Panel C: sex distribution of RHD cases (male n=29; female n=56). Panel D: valve involvement mitral only (n=35); aortic only (n=7); both valves (n=1). Panel E: comparison of echocardiographic RHD prevalence per 1,000 between this study (Dhanusha 2024: 18.7 per 1,000) and the only prior echocardiographic Nepal study (Sunsari 2016: 10.2 per 1,000; Shrestha et al. JAMA Cardiol. 2016;1(1):89–96). RHD = rheumatic heart disease. WHF = World Heart Federation 2012 criteria.

Our prevalence of 18.7 per 1,000 (95% CI 15.1–23.0) places Dhanusha firmly among the high-endemic regions of the world. For comparison, the pooled South Asian estimate from the 2014 Rothenbühler meta-analysis was 28 per 1,000 (95% CI 17–50) [5], the Indian RHEUMATIC study documented 20.4 per 1,000 school children [18], and echocardiographic estimates from sub-Saharan Africa range from 14.2 per 1,000 in Uganda to 30.4 per 1,000 in a combined South African and Ethiopian cohort [19]. In Latin America, a Nicaraguan study found 21.0 per 1,000 [20] and combined Egyptian and Tanzanian estimates exceeded 30 per 1,000 [21]. Our prevalence therefore falls within the range observed in other high-burden regions and is substantially higher than all earlier Nepalese reports, which documented 0.9–7.2 per 1,000 using auscultation-based surveys [10,15,17].

The near-doubling of prevalence relative to Sunsari may be related to Dhanusha’s more disadvantaged socioeconomic and demographic context. Rheumatic heart disease is consistently more common in populations with lower income, rural residence, crowding and limited access to primary healthcare [22–24]. Dhanusha’s higher population density, predominantly agricultural and remittance-dependent economy, and comparatively weaker health infrastructure contrast with Sunsari, which includes the urban centre of Dharan and a tertiary hospital, and could contribute to the higher burden observed. Ethnic differences between the predominantly Madhesi Dhanusha population and the more mixed Sunsari population may also contribute, as inter-ethnic variation in host genetic susceptibility to post-streptococcal autoimmunity and RHD has been demonstrated across South Asian and other populations [25–27].

The age-prevalence distribution in Dhanusha differed markedly from the near-linear increase seen in Sunsari, where RHD prevalence rose from 5.5 per 1,000 at age 5 to 16.0 per 1,000 at age 15 [14]. In Dhanusha, even children aged 6–9 years already carried a substantial burden of 16.0 per 1,000 (males 14.8; females 16.7 per 1,000), with little further rise in older age groups. Comparable high early-age or age-uniform patterns have been described in hyperendemic settings such as Ethiopia and other sub-Saharan African and Oceania cohorts [6,28–30]. If corroborated by future studies, this would argue that in areas sharing Dhanusha’s transmission profile, echocardiographic screening should begin at school entry (≈6 years) rather than prioritizing older children, in contrast to lower-prevalence, more age-progressive regions such as Senegal, where extending screening into later adolescence yielded higher detection rates [31,32].

The predominance of isolated mitral valve disease (41.2%) is consistent with the well-established natural history of rheumatic valvulitis. Pathological MR was present in 89.4% of RHD cases, with peak MR velocity ≥3 m/s and a pansystolic jet each documented in 94.7% of MR-positive cases, confirming the specificity of the WHF 2012 criteria. AML thickening ≥3 mm was present in 42.4% of cases, a morphological feature associated with a higher risk of progression to definite RHD. Aortic involvement was less common (10.6% with AR; 8.2% isolated aortic disease), and combined mitral and aortic disease occurred in only 1.2% of cases. These proportions match those from earlier WHF-based studies in South Asia and sub-Saharan Africa[5,18], and suggest that most cases are early-stage disease, when secondary antibiotic prophylaxis is most effective.

Crucially, none of the echocardiographic or clinical parameters (S1 Table) differed significantly between males and females among the 85 RHD-positive children. This indicates that the sex difference in this cohort operates at the level of susceptibility and disease acquisition—not at the level of disease character or severity once RHD is established.

The association with female sex (OR 1.60, 95% CI 1.02–2.53) was robust in all sensitivity analyses and agrees with findings from the global RHD literature. In Sunsari, female sex carried an OR of 1.86 (95% CI 1.05–3.29) [9]. In Senegal, higher echocardiographic RHD prevalence among girls was documented across all age bands [33]. In India, female sex was among the strongest predictors in the RHEUMATIC study [18]. A meta-analysis of 58 echocardiographic screening studies found lower RHD prevalence in males (RR 0.70 vs females) [34]. Among school-age children, where differential reproductive exposure to streptococci cannot account for the gap, the most biologically plausible mechanism is sex differences in adaptive immune responses: females mount more vigorous T-cell-mediated autoimmune reactivity to streptococcal M-protein epitopes and produce higher titres of anti-cardiac cross-reactive antibodies than males [35–37].

The public health consequence of this female excess extends well beyond childhood. Girls who develop subclinical mitral valve disease during school years carry that disease into reproductive life, where rheumatic mitral stenosis in pregnancy is associated with maternal mortality rates of up to 20% in resource-limited settings [36,38]. In this cohort, 56 of 85 RHD cases (65.9%) were girls. Targeting screening and secondary prophylaxis programmes to girls—particularly those aged 10–14 years at the peak of the prevalence curve—is therefore both clinically necessary and operationally efficient.

### The ICC: school as an independent determinant of RHD risk

The ICC of 0.19 after accounting for sex and age indicates that the school a child attends explains approximately one-fifth of the total variance in their RHD risk, a contribution comparable in magnitude to that of biological sex itself. Several mechanisms plausibly underlie this school-level clustering. Schools act as natural catchment areas for geographically proximate households; structural conditions that promote GAS transmission—household crowding, poor ventilation, limited access to antibiotics for pharyngitis—are therefore shared among children attending the same school [39]. School health services, including the availability of benzathine penicillin G for ARF prophylaxis and health education about streptococcal sore throat, may differ substantially between institutions [40]. Unmeasured school-level variables such as classroom density and hygiene infrastructure may each contribute independently.

The estimated design effect of approximately 109 indicates substantial clustering, such that the sample of 4,536 children across 8 schools corresponds to a much smaller effective sample size for between-school comparisons. Future cluster-based RHD surveillance studies should consider incorporating the ICC into sample size calculations and may require a larger number of clusters to ensure adequate statistical power.

### Subclinical disease and the diagnostic limits of auscultation

The subclinical fraction of 64.7% and the auscultation sensitivity of 35.3% in this study are consistent with the wider global literature. In Sunsari, subclinical disease was approximately five times more common than manifest disease and auscultation sensitivity was only 17% [9]. In high-risk Indigenous Australian children, auscultation alone captured only a minority of echocardiographic RHD cases [41]. The modestly higher auscultation sensitivity in this Dhanusha cohort (35.3% versus 17% in Sunsari) likely reflects the higher absolute prevalence and a consequently greater proportion of cases with more haemodynamically advanced lesions—a finding with an important implication: even in a high-prevalence setting where auscultation is at its relative best, nearly two-thirds of affected children would remain undiagnosed without echocardiography [9]. Those children would forgo secondary prophylaxis until they present with advanced valvular damage, frequently during pregnancy or when symptomatic heart failure develops in early adulthood. Data from the global REMEDY registry confirm that patients in low-income countries present late, with severe multivalve disease and that the gap in surgical capacity is severe [42].

Given the high RHD prevalence, large school-level ICC, and high proportion of subclinical cases, portable echocardiography should be integrated into routine school health programmes in Dhanusha and similar districts across Nepal. Screening should start at school entry (age 6) given the already high burden detectable in the youngest age group. Because the ICC establishes that schools cluster RHD risk substantially, targeting high-risk schools—rather than universal individual screening—is likely to be the more cost-efficient deployment of limited echocardiographic resources [43]. All identified borderline and definite RHD cases should receive secondary benzathine penicillin G prophylaxis and annual echocardiographic reassessment. The 54 borderline RHD cases identified in this study are currently under active follow-up at SGNHC Janakpur; this longitudinal registry will help determine what proportion of borderline lesions progress to definite disease under real-world prophylaxis conditions. Formal cost-effectiveness analysis of different screening strategies—including optimal number of schools, re-screening intervals and targeted versus universal approaches—is an urgent priority for Nepal’s Ministry of Health and Population.

Strengths of this study include the large sample from a previously unstudied high-burden district, consistent application of WHF 2012 criteria enabling direct international comparison, a rigorous two-cardiologist echocardiographic protocol with consensus-based diagnosis, the use of random-effects modelling specifically appropriate for a small number of clusters and the provision of the first ICC estimate for RHD from Nepal. Limitations are as follows. The small number of schools (n=8) yields wide ICC confidence intervals (0.07–0.44); the design effect estimate should be used cautiously and ideally validated in a larger future study with at least 20–25 schools. The restriction to public schools may modestly overestimate true district-wide prevalence. No formal inter-observer reliability assessment was performed for echocardiographic readings, although all suspicious findings were independently reviewed by a second senior cardiologist before any diagnosis was finalised. The cross-sectional design precludes causal inference and does not allow determination of progression rates from borderline to definite disease, which requires longitudinal follow-up. Finally, the single-district scope limits direct generalisability to other Terai districts or to Nepal’s hill regions.

## Conclusions

RHD affects 18.7 per 1,000 school-going children in Dhanusha district, the highest documented echocardiographic burden in Nepal, with the peak falling among girls aged 10–14 years at 26.0 per 1,000. Two-thirds of cases are subclinical and would be missed by auscultation alone. Female sex is the only independent individual-level predictor and the echocardiographic phenotype of RHD does not differ significantly by sex once disease is established. The school-level ICC of 0.19 reveals that school attended explains as much of a child’s RHD risk as their biological sex, identifying schools as a modifiable structural target for both screening strategy and environmental intervention. Future echocardiographic surveillance studies in Nepal’s Terai must enrol at least 20–25 schools and incorporate this ICC in sample size calculations. Integrating portable echocardiographic screening into routine school health programmes, targeting high-risk schools and prioritising girls from age 6, is both warranted and urgent.

## Data Availability

The data underlying the findings of this study are stored on a secure, encrypted computer system at the Nepal Heart Foundation (NHF). To ensure participant confidentiality and comply with institutional security protocols, access is restricted solely to the Principal Investigator, Prof. Dr. Prakash Raj Regmi (Director of the Nepal RHD Control,Professor of Cardiology, National Academy of Medical Sciences National Program Director, Nepal Heart Foundation President, Nepal Non-communicable Disease Alliance Program and Past President of NHF). Due to these legal and ethical restrictions, the raw data are not publicly available. However, interested, qualified researchers may request access to the data by contacting the Nepal Heart Foundation Central Office at. Data access will be granted upon reasonable request and subject to approval by the NHF Ethics Committee.

## Acknowledgments

The authors extend their gratitude to the principals, teachers, parents, and children of all eight participating schools in Dhanusha district for their cooperation and trust. The Nepal Heart Foundation data management team and field interviewers provided essential logistical support throughout data collection. We thank the clinical staff of the Shahid Gangalal National Heart Centre branch hospital, Janakpur, for ensuring uninterrupted follow-up care of all identified RHD cases. We also acknowledge Mr. Prithutam Bhattarai for their valuable contributions to the study.

## Supporting information

S1 Checklist. Completed STROBE checklist for cross-sectional observational studies with manuscript page numbers for all 22 items. Supplementary Table:

**S1.**
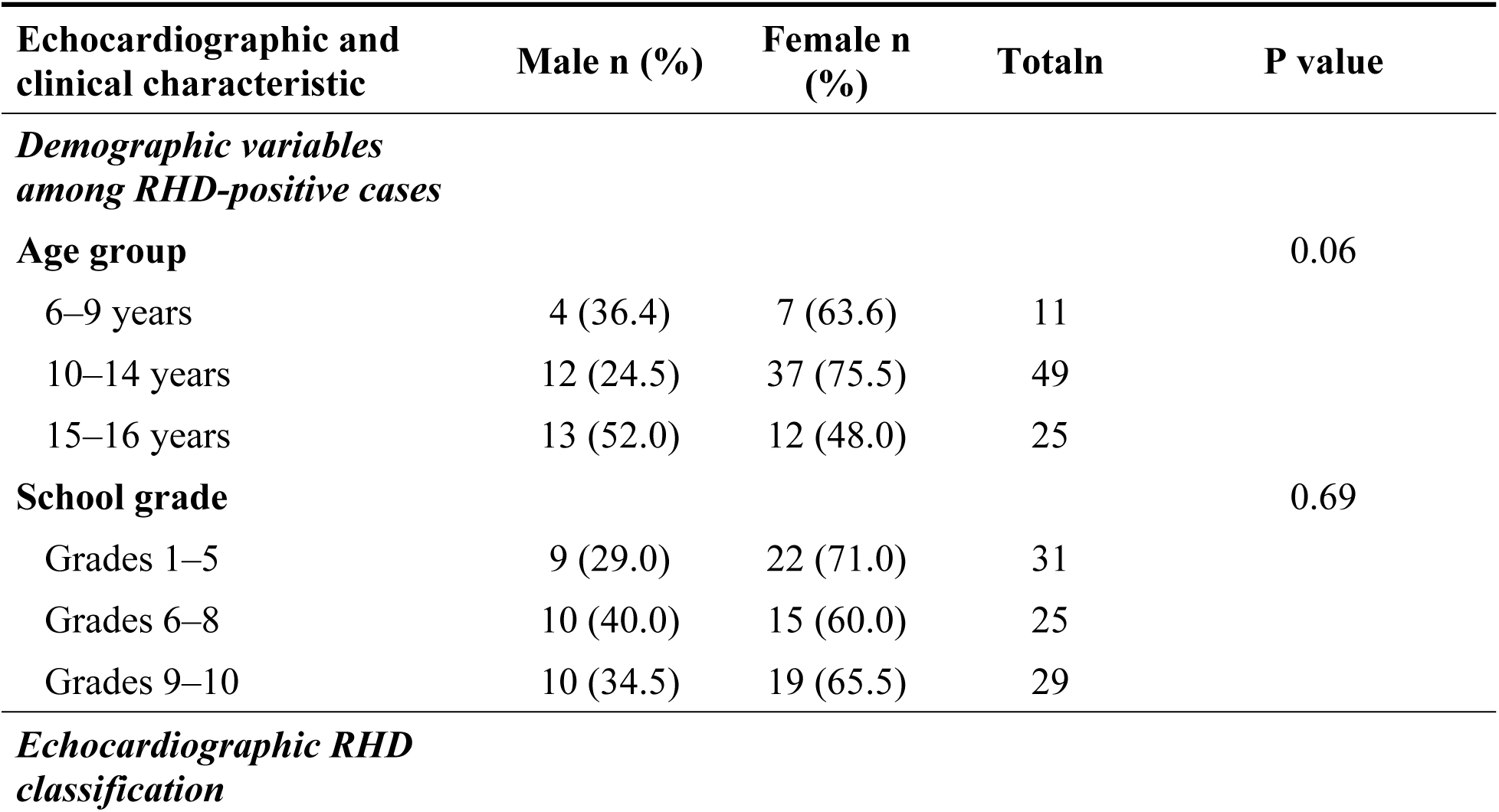
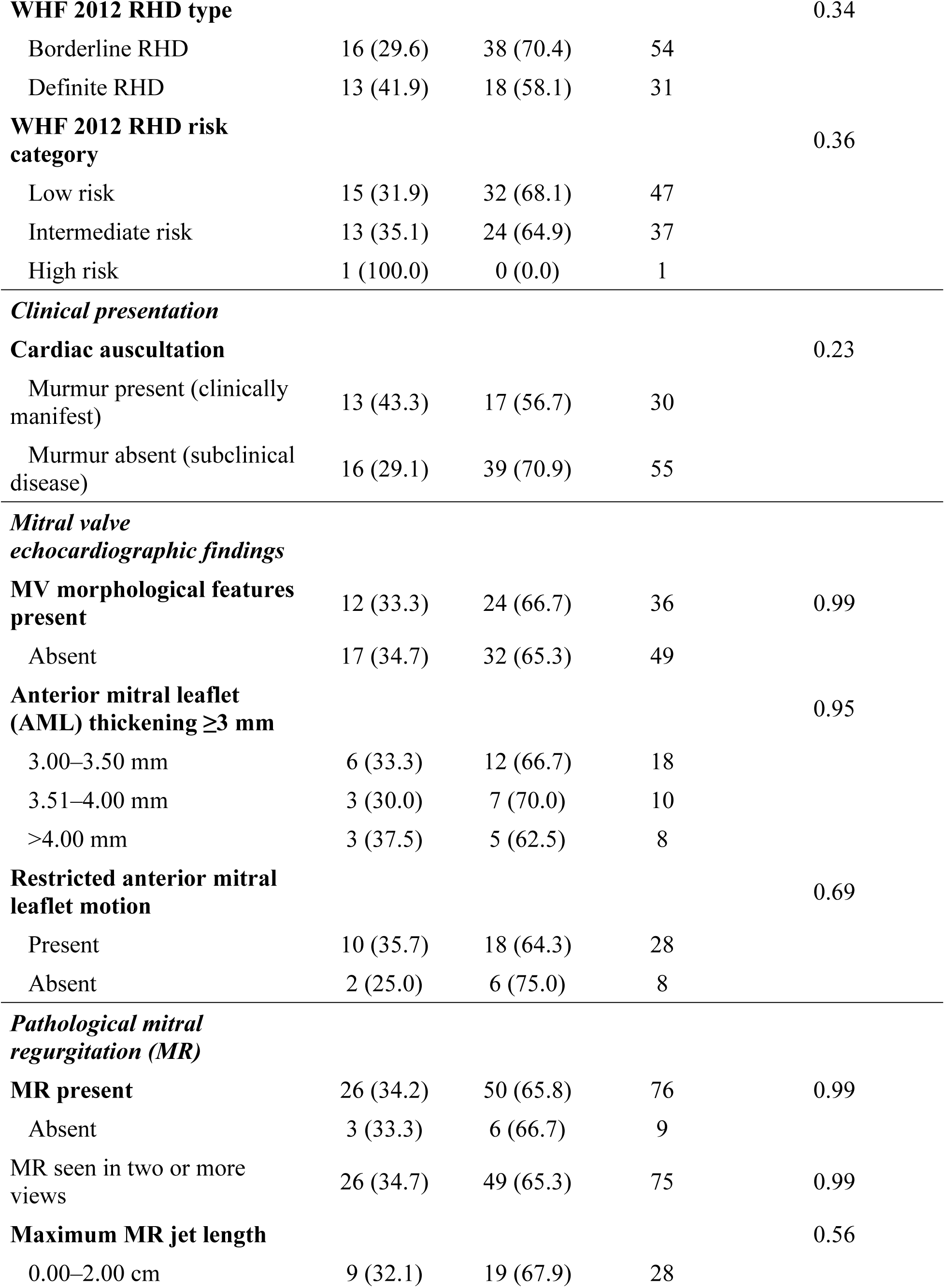
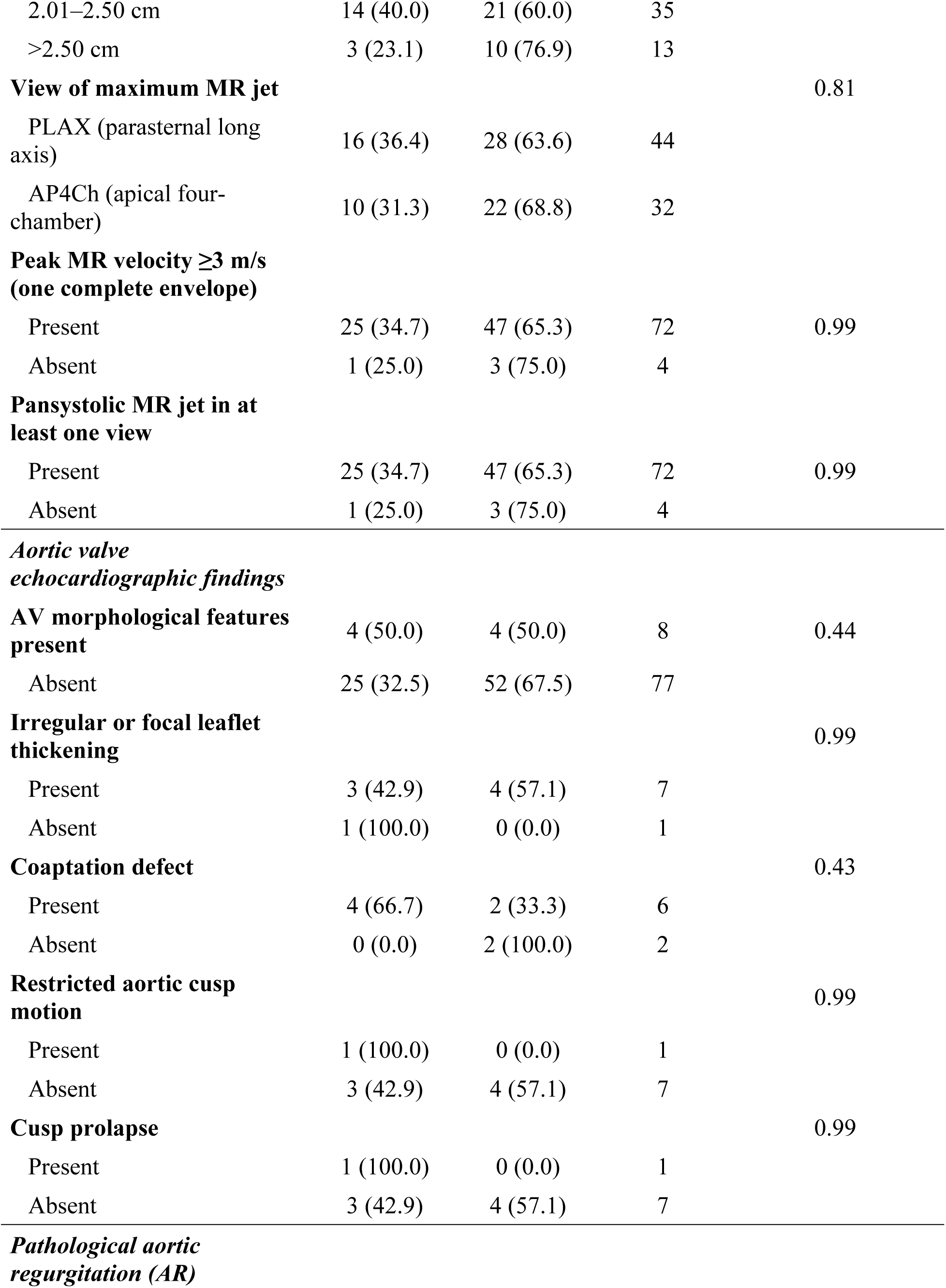
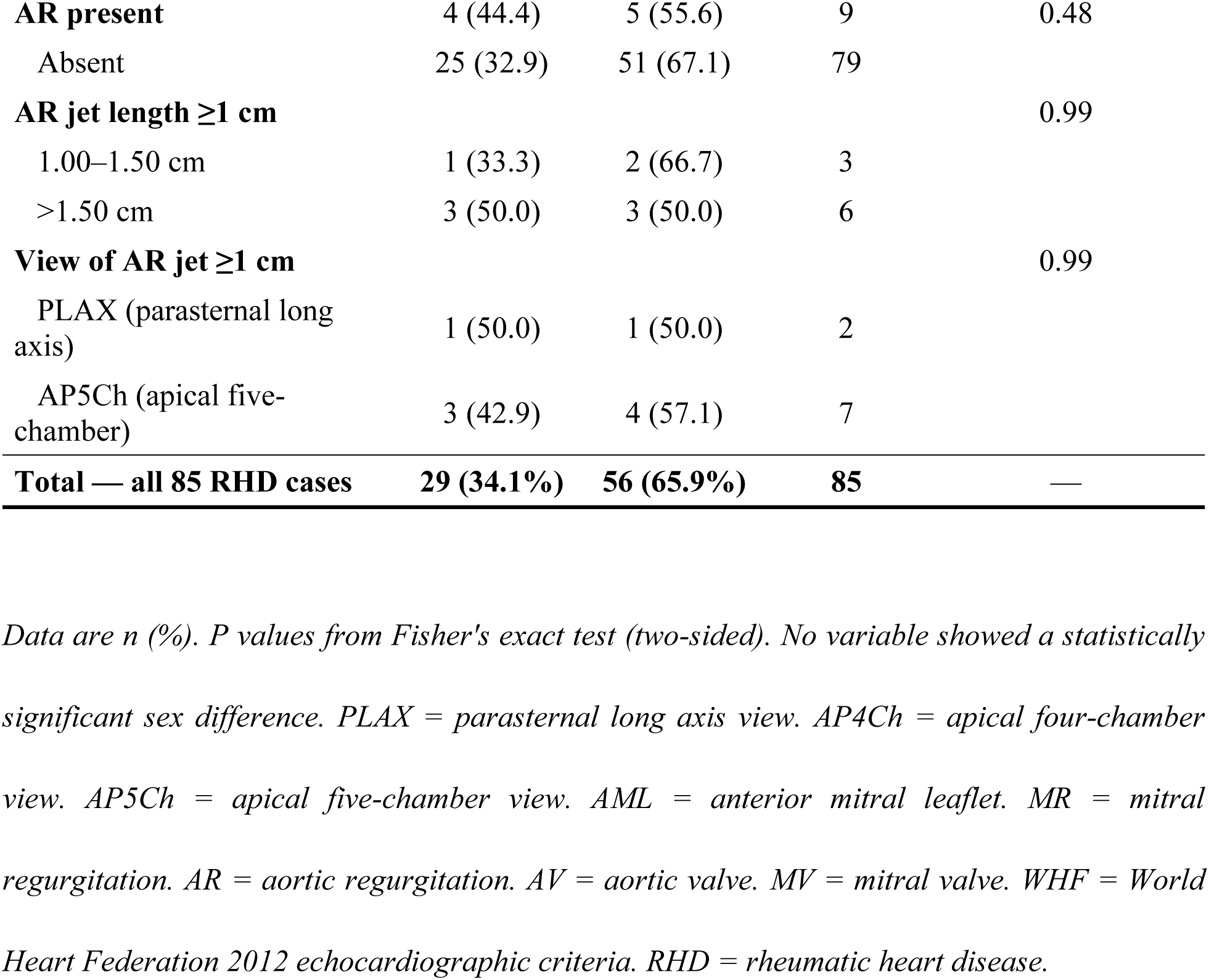
Echocardiographic and clinical findings among the 85 RHD-positive children stratified by sex, Dhanusha district, Nepal, 2023–2024, (n=4,536)

| Echocardiographic and clinical characteristic | Male n (%) | Female n (%) | Totaln | P value |
| --- | --- | --- | --- | --- |
| *Demographic variables among RHD-positive cases* |  |  |  |  |
| **Age group** |  |  |  | 0.06 |
| 6–9 years | 4 (36.4) | 7 (63.6) | 11 |  |
| 10–14 years | 12 (24.5) | 37 (75.5) | 49 |  |
| 15–16 years | 13 (52.0) | 12 (48.0) | 25 |  |
| **School grade** |  |  |  | 0.69 |
| Grades 1–5 | 9 (29.0) | 22 (71.0) | 31 |  |
| Grades 6–8 | 10 (40.0) | 15 (60.0) | 25 |  |
| Grades 9–10 | 10 (34.5) | 19 (65.5) | 29 |  |
| *Echocardiographic RHD classification* |  |  |  |  |
| **WHF 2012 RHD type** |  |  |  | 0.34 |

|  |  |  |  |
| --- | --- | --- | --- |
| Borderline RHD | 16 (29.6) | 38 (70.4) | 54 |
| Definite RHD | 13 (41.9) | 18 (58.1) | 31 |
| **WHF 2012 RHD risk category** |  |  |  | 0.36 |

|  |  |  |  |
| --- | --- | --- | --- |
| Low risk | 15 (31.9) | 32 (68.1) | 47 |
| Intermediate risk | 13 (35.1) | 24 (64.9) | 37 |
| High risk | 1 (100.0) | 0 (0.0) | 1 |
| ***Clinical presentation*** |  |  |  |  |
| **Cardiac auscultation** |  |  |  | 0.23 |

|  |  |  |  |
| --- | --- | --- | --- |
| Murmur present (clinically manifest) | 13 (43.3) | 17 (56.7) | 30 |
| Murmur absent (subclinical disease) | 16 (29.1) | 39 (70.9) | 55 |
| **MV morphological features present** | 12 (33.3) | 24 (66.7) | 36 | 0.99 |

|  |  |  |  |
| --- | --- | --- | --- |
| Absent | 17 (34.7) | 32 (65.3) | 49 |
| **Anterior mitral leaflet (AML) thickening $\geq 3$ mm** |  |  |  | 0.95 |

|  |  |  |  |
| --- | --- | --- | --- |
| 3.00–3.50 mm | 6 (33.3) | 12 (66.7) | 18 |
| 3.51–4.00 mm | 3 (30.0) | 7 (70.0) | 10 |
| >4.00 mm | 3 (37.5) | 5 (62.5) | 8 |
| **Restricted anterior mitral leaflet motion** |  |  |  | 0.69 |

|  |  |  |  |
| --- | --- | --- | --- |
| Present | 10 (35.7) | 18 (64.3) | 28 |
| Absent | 2 (25.0) | 6 (75.0) | 8 |
| ***Pathological mitral regurgitation (MR)*** |  |  |  |  |
| **MR present** | 26 (34.2) | 50 (65.8) | 76 | 0.99 |

|  |  |  |  |
| --- | --- | --- | --- |
| Absent | 3 (33.3) | 6 (66.7) | 9 |
| MR seen in two or more views | 26 (34.7) | 49 (65.3) | 75 | 0.99 |
| **Maximum MR jet length** |  |  |  | 0.56 |

|  |  |  |  |
| --- | --- | --- | --- |
| 0.00–2.00 cm | 9 (32.1) | 19 (67.9) | 28 |
| 2.01–2.50 cm | 14 (40.0) | 21 (60.0) | 35 |  |
| >2.50 cm | 3 (23.1) | 10 (76.9) | 13 |  |
| **View of maximum MR jet** |  |  |  | 0.81 |
| PLAX (parasternal long axis) | 16 (36.4) | 28 (63.6) | 44 |  |
| AP4Ch (apical four-chamber) | 10 (31.3) | 22 (68.8) | 32 |  |
| **Peak MR velocity $\geq 3$ m/s (one complete envelope)** |  |  |  |  |
| Present | 25 (34.7) | 47 (65.3) | 72 | 0.99 |
| Absent | 1 (25.0) | 3 (75.0) | 4 |  |
| **Pansystolic MR jet in at least one view** |  |  |  |  |
| Present | 25 (34.7) | 47 (65.3) | 72 | 0.99 |
| Absent | 1 (25.0) | 3 (75.0) | 4 |  |
| ***Aortic valve echocardiographic findings*** |  |  |  |  |
| **AV morphological features present** | 4 (50.0) | 4 (50.0) | 8 | 0.44 |
| Absent | 25 (32.5) | 52 (67.5) | 77 |  |
| **Irregular or focal leaflet thickening** |  |  |  | 0.99 |
| Present | 3 (42.9) | 4 (57.1) | 7 |  |
| Absent | 1 (100.0) | 0 (0.0) | 1 |  |
| **Coaptation defect** |  |  |  | 0.43 |
| Present | 4 (66.7) | 2 (33.3) | 6 |  |
| Absent | 0 (0.0) | 2 (100.0) | 2 |  |
| **Restricted aortic cusp motion** |  |  |  | 0.99 |
| Present | 1 (100.0) | 0 (0.0) | 1 |  |
| Absent | 3 (42.9) | 4 (57.1) | 7 |  |
| **Cusp prolapse** |  |  |  | 0.99 |
| Present | 1 (100.0) | 0 (0.0) | 1 |  |
| Absent | 3 (42.9) | 4 (57.1) | 7 |  |
| ***Pathological aortic regurgitation (AR)*** |  |  |  |  |
| **AR present** | 4 (44.4) | 5 (55.6) | 9 | 0.48 |
| Absent | 25 (32.9) | 51 (67.1) | 79 |  |
| **AR jet length $\geq 1$ cm** |  |  |  | 0.99 |
| 1.00–1.50 cm | 1 (33.3) | 2 (66.7) | 3 |  |
| >1.50 cm | 3 (50.0) | 3 (50.0) | 6 |  |
| **View of AR jet $\geq 1$ cm** |  |  |  | 0.99 |
| PLAX (parasternal long axis) | 1 (50.0) | 1 (50.0) | 2 |  |
| AP5Ch (apical five-chamber) | 3 (42.9) | 4 (57.1) | 7 |  |
| **Total — all 85 RHD cases** | **29 (34.1%)** | **56 (65.9%)** | **85** | **—** |

Data are n (%). P values from Fisher's exact test (two-sided). No variable showed a statistically significant sex difference. PLAX = parasternal long axis view. AP4Ch = apical four-chamber view. AP5Ch = apical five-chamber view. AML = anterior mitral leaflet. MR = mitral regurgitation. AR = aortic regurgitation. AV = aortic valve. MV = mitral valve. WHF = World Heart Federation 2012 echocardiographic criteria. RHD = rheumatic heart disease.

**S2.** Rheumatic heart disease (RHD)-related symptoms reported in the past 12 months and their association with echocardiographic RHD status, (n=4,536)

| Symptom (past 12 months) | Reported<br>n (%) | No RHD<br>n (%) | RHD<br>n (%) | P value |
| --- | --- | --- | --- | --- |
| **Throat pain or tonsillitis with fever** |  |  |  |  |
| Yes | 1,995 (44.0) | 1,965 (98.5) | 30 (1.5) | 0.10 |
| No | 2,541 (56.0) | 2,486 (97.8) | 55 (2.2) |  |
| **Fever with joint pain in hands, shoulders, hips or legs** |  |  |  |  |
| Yes | 1,176 (25.9) | 1,156 (98.3) | 20 (1.7) | 0.71 |
| No | 3,360 (74.1) | 3,295 (98.1) | 65 (1.9) |  |
| **Involuntary jerky movements of the body (chorea)** |  |  |  |  |
| Yes | 965 (21.3) | 948 (98.2) | 17 (1.8) | 0.89 |
| No | 3,571 (78.7) | 3,503 (98.1) | 68 (1.9) |  |
| **Any of the three symptoms above (at any time)** |  |  |  |  |
| Yes | 2,586 (57.0) | 2,548 (98.5) | 38 (1.5) | 0.03 |
| No‡ | 1,950 (43.0) | 1,903 (97.6) | 47 (2.4) |  |
| **All three symptoms present simultaneously** |  |  |  |  |
| Yes | 449 (9.9) | 440 (98.0) | 9 (2.0) | 0.85 |
| No | 4,087 (90.1) | 4,011 (98.1) | 76 (1.9) |  |

*Data are n (%). P values from chi-squared or Fisher's exact test. ‡Children with no symptom history had a higher RHD prevalence (2.4%) than those with any symptom history (1.5%; P=0.03), reflecting the predominantly subclinical nature of echocardiographically detected disease. RHD = rheumatic heart disease.*

